# Electrolyte Abnormalities and Clinical Outcomes in Heart Failure Patients: A Retrospective Cohort Study

**DOI:** 10.1101/2025.11.11.25340038

**Authors:** Shankar Biswas, Bilal Qammar, Yashasvi Srivastava, Chaudhari Mahendrakumar Achlaram, Mirza Mohammad Ali Baig, Muhammad Umar, Ibrahim Manzoor, Bilal Tariq, Elangovan Krishnan, Jamal S. Rana

## Abstract

**Background:** Electrolyte abnormalities commonly complicate heart failure management, yet their prognostic significance and optimal monitoring strategies remain incompletely characterized. We examined the prevalence, temporal patterns, and clinical outcomes associated with electrolyte abnormalities in hospitalized heart failure patients.

**Methods:** Retrospective cohort study of 30,678 heart failure patients (80,408 admissions) from the MIMIC-IV database (2008-2022). Clinically significant electrolyte abnormalities (CSEA) were defined as documented potassium <3.5 or >5.0 mEq/L and sodium <135 or >145 mEq/L during hospitalization. Primary outcomes were 30-day all-cause readmission and mortality. We developed a clinical risk score incorporating electrolyte abnormalities and examined temporal electrolyte patterns.

**Results:** CSEA occurred in 705 patients (2.3%), with 96.7% achieving normalization by discharge. Patients with abnormalities had paradoxically lower 30-day readmission (3.9% vs 15.3%; adjusted HR 0.29, 95% CI 0.21-0.40) but substantially higher mortality (15.4% vs 7.0%; adjusted HR 1.93, 95% CI 1.57-2.38), reflecting competing mortality risk. Individual electrolyte abnormalities independently predicted mortality: hypokalemia HR 1.45 (1.28-1.65), hyperkalemia HR 1.67 (1.48-1.89), hyponatremia HR 1.34 (1.19-1.51). Temporal analysis revealed biphasic patterns—initial correction followed by recurrence—identified the highest-risk subset (composite event rates 27.6% for potassium, 28.5% for sodium). A five-variable risk score (electrolyte abnormality, age ≥75, chronic kidney disease, multiple admissions, coronary artery disease) achieved C-statistic 0.595 with 2.34-fold risk discrimination across categories.

**Conclusions:** CSEA independently predict mortality in heart failure patients despite paradoxically lower readmission rates due to competing risks. Biphasic electrolyte patterns identify particularly high-risk patients. Simple risk stratification using routinely collected electrolyte data may enhance post-discharge risk prediction and inform targeted monitoring strategies.

## INTRODUCTION

Heart failure (HF) occurs when structural or functional abnormalities of the heart lead to a syndrome whereby the ventricles are unable to fill with or eject blood. It culminates the majority of cardiovascular diseases, including ischemic heart disease, hypertension, valve disease, and cardiomyopathies [1]. It affects over 64 million individuals worldwide, and the prevalence increases with population aging, greater post-myocardial infarction survival, and increased recognition of chronic heart conditions [2]. Despite the greater use of pharmacological therapies and heart devices, the failure of a heart to pump blood adequately results in a high degree of morbidity, frequent hospitalizations, and considerable economic costs [3]. The disease is highly lethal, and five-year survival is often better than common cancers like breast and prostate cancer. Heart failure (HF) is defined as the inability of the heart to function properly as a pump, and therefore not able to maintain adequate perfusion to the tissues under various physiological demands [4]. More than 64.3 million individuals globally suffer from HF [5]. With increases in life expectancy, this figure will likely continue to grow [6]. The combination of greater than 30% mortality per year, high economic costs, and the adverse effects of HF on the quality of life of the patients make it a major global health issue. The heterogeneous pathophysiological characteristics of HF justify the numerous different classifications, as it is the only condition in medicine to be classified into 5 different diseases [7]. The maintenance of normal myocardial function and systemic hemodynamic equilibrium is influenced significantly by electrolyte homeostasis. The electrolytes sodium (Na⁺), potassium (K⁺), chloride (Cl⁻), calcium (Ca²⁺), and magnesium (Mg²⁺), and to lesser extent the skeletal muscles, and all the cardiac muscles, control electrical excitability, contractility, and various enzymatic activities [8]. In patients with HF, the electrolyte disturbances are secondary to the overlapping mechanisms of neurohormonal activation, renal failure, diuretic and neurohormonal blocker medications. These electrolyte disturbances go beyond being a laboratory curiosity, as they are closely associated with the severity of the disease and are key factors influencing the course and outcome of the hospital stay, as well as the hospital admission [9].

Patients with heart failure (HF) frequently contend with electrolyte abnormalities (EAs), which result from HF’s own complications, as well as the various therapeutic agents. Even in those healthcare facilities which incorporate routine screenings, EAs are often undocumented and underappreciated [10]. For example, hypokalaemia and hypomagnesemia account for half of all sudden deaths from HF and associated ventricular arrhythmias [11]. Meanwhile, elevated potassium levels are associated with an increased risk of all-cause mortality after hospitalization for HF [12]. In hospitalised patients with HF, hypochloremia correlated with an increased risk of mortality, need for more inotropic agents, and poor response to diuretics [13]. Complications of HF such as atrial fibrillation and myocardial ischemia are associated with the troika of unbalanced magnesium, phosphate, and calcium [14]. Hyponatremia, defined as a serum sodium concentration below 135 mmol/L, enjoys the dubious distinction of being the most often seen electrolyte anomaly in patients with HF. It signifies advanced HF and is associated with poor outcomes. The mechanism for this includes the non-osmotic capture of the arginine vasopressin, which causes free water retention causing dilutional hyponatremia [15]. The hyponatremia was associated with increased in-hospital mortality, prolonged hospital stays, greater readmission rates, and mortality after discharge in studies like the OPTIMIZE-HF registry. On the other hand, hypernatremia may occur due to excessive use of diuretics, osmotic diuresis, and controlling it may also indicate poor prognosis [16]. Lastly, the imbalance of potassium is also an important outcome predictor of HF. Hypokalemia is sometimes due to the use of loop and thiazide diuretics, hyperaldosteronism, or poor dietary intake. Hypokalemia poses an even greater threat due to the increased risk of ventricular arrhythmias, sudden cardiac death, and digitalis toxicity [17]. Conversely, hyperkalemia is more common among those with renal dysfunction taking RAAS inhibitors or potassium-sparing diuretics. Mortality risk increases with both. Hence, the risk associated with an extreme of either level of the potassium and the need for close monitoring [18].

Associational evidence linking poor outcomes to hypokalaemia in patients with HF, including elevated all-cause mortality, has been demonstrated in large observational studies as well as clinical trials like TOPCAT and RALES [19–21]. In addition, studies show that hypokalaemia and poor prognoses in HF do not significantly differ with respect to LVEF [31]. In EMPHASIS-HF, eplerenone significantly decreased hypokalaemia incidence (HR 0.69; p < 0.001), and 26% of the cardiovascular benefits of eplerenone were attributed to the correction of hypokalaemia [22]. Conversely, a study involving patients post cardiac surgery found that more relaxed potassium control (≥3.6 mEq/L) in comparison to tight control (≥4.5 mEq/L) control was not associated with increased rates of atrial fibrillation and other HF aggravating arrhythmias, or death during the hospitalization [23]. Given these circumstances, we designed a retrospective cohort study to assess the extent and range of electrolyte abnormalities in patients with heart failure and their association with important clinical outcomes, including in-hospital mortality, length of stay, and rates of readmission. This study seeks to illustrate the associations that will demonstrate the importance of considering the early recognition of electrolyte imbalance as an integral part of heart failure management and recognizing the bounds identifying the need for immediate treatment.

## METHODS

### Study Design and Data Source

We conducted a retrospective cohort study using the Medical Information Mart for Intensive Care IV version 3.1 (MIMIC-IV v3.1) database, a large, publicly available, de-identified critical care database from Beth Israel Deaconess Medical Center in Boston, Massachusetts [24, 25]. The database contains comprehensive clinical data from 2008 to 2022, including demographics, vital signs, laboratory values, medications, diagnoses, procedures, and outcomes for intensive care unit patients. Data access was obtained through completion of the Collaborative Institutional Training Initiative (CITI) program and execution of a data use agreement. As MIMIC-IV contains only de-identified data, this study was exempt from institutional review board approval. This study strictly adhered to the Strengthening the Reporting of Observational Studies in Epidemiology (STROBE) guidelines.

### Study Population

We identified all adult patients (age ≥18 years) with a primary diagnosis of heart failure during the study period using International Classification of Diseases, Ninth and Tenth Revision (ICD-9 and ICD-10) codes [26]. From an initial cohort of 80,611 heart failure admissions, we excluded 203 admissions with incomplete demographic data, resulting in a final analytical cohort of 80,408 admissions representing 30,678 unique patients. For patient-level analyses, we focused on the last recorded admission for each patient when outcome data were available (n=28,154, 91.8%). Patients contributed a mean of 2.6 ± 3.7 admissions (median 1, IQR 1-3; range 1-164), with 13,944 patients (45.5%) having multiple heart failure admissions during the study period.

### Variable Definitions Electrolyte Abnormalities

The primary exposure was the presence of electrolyte abnormalities during hospitalization. We defined electrolyte abnormalities as: hypokalemia (serum potassium <3.5 mEq/L), hyperkalemia (serum potassium >5.0 mEq/L), hyponatremia (serum sodium <135 mEq/L), and hypernatremia (serum sodium >145 mEq/L). Electrolyte measurements were obtained from routine laboratory testing throughout the hospitalization. The median number of potassium and sodium measurements per admission was 6 (IQR 3-10).

For the primary outcome analyses, we classified patients as having clinically significant electrolyte abnormalities (CSEA) if they had documented abnormal values that were flagged in the patient record during hospitalization (n=705, 2.3% of cohort). This definition captured patients with abnormalities of sufficient severity and duration to warrant clinical documentation and intervention. For baseline characteristics comparison, we separately identified patients with any abnormal electrolyte values at the time of discharge (n=1,024, 3.3% of cohort), which included both clinically significant abnormalities and milder derangements that persisted until discharge. The discharge electrolyte status was used for baseline comparisons to characterize the full spectrum of electrolyte abnormalities in the cohort, while the clinically significant abnormalities during hospitalization were used as the exposure for all outcome analyses.

In sensitivity analyses, we also examined outcomes using alternative definitions including any abnormal value during hospitalization and varying threshold criteria (potassium: 3.0-5.5 mEq/L [conservative], 3.5-5.0 mEq/L [standard], 3.8-4.5 mEq/L [strict]; sodium: 130-150 mEq/L [mild], 135-145 mEq/L [standard], 138-142 mEq/L [strict]).

We additionally characterized temporal electrolyte patterns by examining serial measurements throughout hospitalization. For both potassium and sodium, we classified patients into trajectory patterns: (1) stable normal (maintained normal values throughout), (2) corrected (initial abnormality that was corrected and maintained), (3) developed (normal on admission with subsequent abnormality), and (4) biphasic (abnormality with correction followed by recurrence). Discharge electrolyte status was determined from the final laboratory values obtained before hospital discharge.

### Demographics and Comorbidities

We extracted demographic data including age, sex, race/ethnicity, and insurance type from admission records. Comorbidities were identified using ICD-9 and ICD-10 diagnosis codes and classified according to the Elixhauser comorbidity classification system [27]. Specific comorbidities assessed included hypertension, coronary artery disease, chronic kidney disease, anemia, chronic obstructive pulmonary disease, atrial fibrillation, diabetes mellitus, depression, malignancy, and obesity. We calculated the Charlson Comorbidity Index for each patient as a measure of overall comorbidity burden [28].

### Medications

Heart failure medications were identified from pharmacy records and medication administration records. We assessed use of guideline-directed medical therapy [29] including angiotensin-converting enzyme inhibitors, angiotensin receptor blockers, beta-blockers, diuretics, and aldosterone antagonists, as well as other relevant medications including anticoagulants, statins, antiplatelets, digoxin, and insulin. Medication data were analyzed both at the admission level (medications during a specific hospitalization) and at the patient level (ever prescribed during any hospitalization).

### Clinical Characteristics

Length of stay was calculated as the time from admission to discharge in days. For patients with multiple admissions, we recorded the total number of heart failure admissions during the study period. Laboratory values including serum creatinine, blood urea nitrogen, hemoglobin, and when available, B-type natriuretic peptide, were extracted from laboratory records.

### Outcome Definitions

The primary outcomes were 30-day all-cause readmission and 30-day mortality after discharge. We also examined a composite outcome of 30-day readmission or mortality. Secondary outcomes included heart failure-specific 30-day readmission (readmission with primary diagnosis of heart failure), 7-day all-cause and heart failure-specific readmission, in-hospital mortality, and time to first readmission. Readmission and mortality data were obtained from administrative records and death registries. Outcome data were available for 95% of the cohort.

### Statistical Analysis Descriptive Analysis

We characterized the cohort using descriptive statistics, presenting continuous variables as mean ± standard deviation for normally distributed data or median with interquartile range for skewed data, and categorical variables as number and percentage. We compared baseline characteristics between patients with any abnormal electrolytes at discharge (n=1,024, 3.3%) and those with normal electrolytes at discharge (n=29,654, 96.7%) using chi-square tests or Fisher’s exact test for categorical variables and t-tests or Mann-Whitney U tests for continuous variables, as appropriate. This comparison provided a comprehensive characterization of patients with electrolyte abnormalities across the severity spectrum. Statistical significance was defined as a two-tailed p-value <0.05.

### Unadjusted Analysis

We calculated unadjusted odds ratios with 95% confidence intervals for the association between CSEA (n=705) and each outcome using logistic regression. We compared outcome rates between patients with normal electrolytes and those with clinically significant abnormalities using chi-square tests.

### Multivariable Analysis

We used Cox proportional hazards regression [30] to examine the independent association between CSEA and time to each outcome (30-day all-cause readmission, heart failure-specific readmission, and mortality), adjusting for potential confounders including age (continuous), sex, chronic kidney disease, diabetes mellitus, and coronary artery disease. We assessed the proportional hazards assumption using Schoenfeld residuals and log-log plots. Model discrimination was assessed using Harrell’s concordance index (C-index). Results are reported as hazard ratios with 95% confidence intervals.

### Risk Score Development

We developed a clinical risk score to predict the 30-day composite outcome (readmission or mortality) using multivariable logistic regression. Variables were selected based on clinical relevance, statistical significance in univariate analysis, availability at the time of discharge, and ease of clinical use. Point values were assigned to each component based on beta coefficients from the logistic regression model, rounded to integers for clinical practicality. The final risk score included: any electrolyte abnormality (2 points), age ≥75 years (1 point), chronic kidney disease (1 point), multiple heart failure admissions (1 point), and coronary artery disease (1 point), for a total possible score of 0-7 points.

We categorized patients into risk groups: low risk (score 0), moderate risk (score 1-2), high risk (score 3-4), and very high risk (score ≥5). We assessed risk score performance using the C-statistic with 95% confidence interval, and calibration using the Hosmer-Lemeshow goodness-of-fit test and calibration plots. We calculated net reclassification improvement compared to clinical judgment alone and quantified risk discrimination by comparing event rates across risk categories.

### Subgroup and Sensitivity Analyses

We performed pre-specified subgroup analyses to examine effect modification by age (<65, 65-74, 75-84, ≥85 years), sex, chronic kidney disease status and severity, diabetes, and presence of multiple admissions. We tested for statistical interaction by including interaction terms in multivariable models. Results are presented stratified by subgroup with forest plots for visualization.

We conducted sensitivity analyses using alternative electrolyte threshold definitions and varying outcome definitions (readmission only, mortality only, composite) to assess the robustness of our findings. We also examined associations separately at the admission level to account for patients with multiple hospitalizations.

### Temporal Pattern and Medication Analyses

We compared outcomes across electrolyte trajectory patterns using chi-square tests and calculated event rates for each pattern. We assessed guideline-directed medical therapy use and tolerance by comparing medication prescription rates between patients with normal and abnormal electrolytes and examined associations between medication use and electrolyte normalization rates.

### Quality Metrics

We calculated quality performance indicators including rates of electrolyte normalization at discharge, length of stay by electrolyte status, and readmission rates stratified by discharge electrolyte status. We compared current performance to quality benchmarks and identified gaps and opportunities for quality improvement.

### Data Management and Software

Data extraction and processing followed a systematic pipeline including covariate extraction, electrolyte aggregation, outcome linkage, and patient-level dataset creation. We documented missing data patterns and assessed data completeness for all key variables. All statistical analyses were performed using Python and SQL. We conducted complete case analyses without imputation for missing data. For patients with multiple admissions, the primary analysis used the last admission with available outcome data. All tests were two-tailed with alpha=0.05.

### Sample Size and Statistical Power

With 30,678 patients including 5,818 composite outcome events (19.0%) and 705 patients (2.3%) with CSEA, our study had >90% power to detect clinically meaningful effect sizes including odds ratios ≥1.5 for mortality and ≤0.7 for readmission at a two-sided significance level of 0.05.

## RESULTS

### Study Population

Our study included 30,678 unique heart failure patients representing 80,408 hospital admissions during the study period. The mean age was 74.1 ± 13.4 years and 14,325 (46.7%) were female. Detailed baseline characteristics for the entire cohort are provided in Supplementary Table S1. Nearly half of patients (13,944, 45.5%) had multiple heart failure admissions, with a mean of 2.6 ± 3.7 admissions per patient. Outcome data were available for 28,154 patients (91.8%) **(Figure 1).**

**Figure 1.**
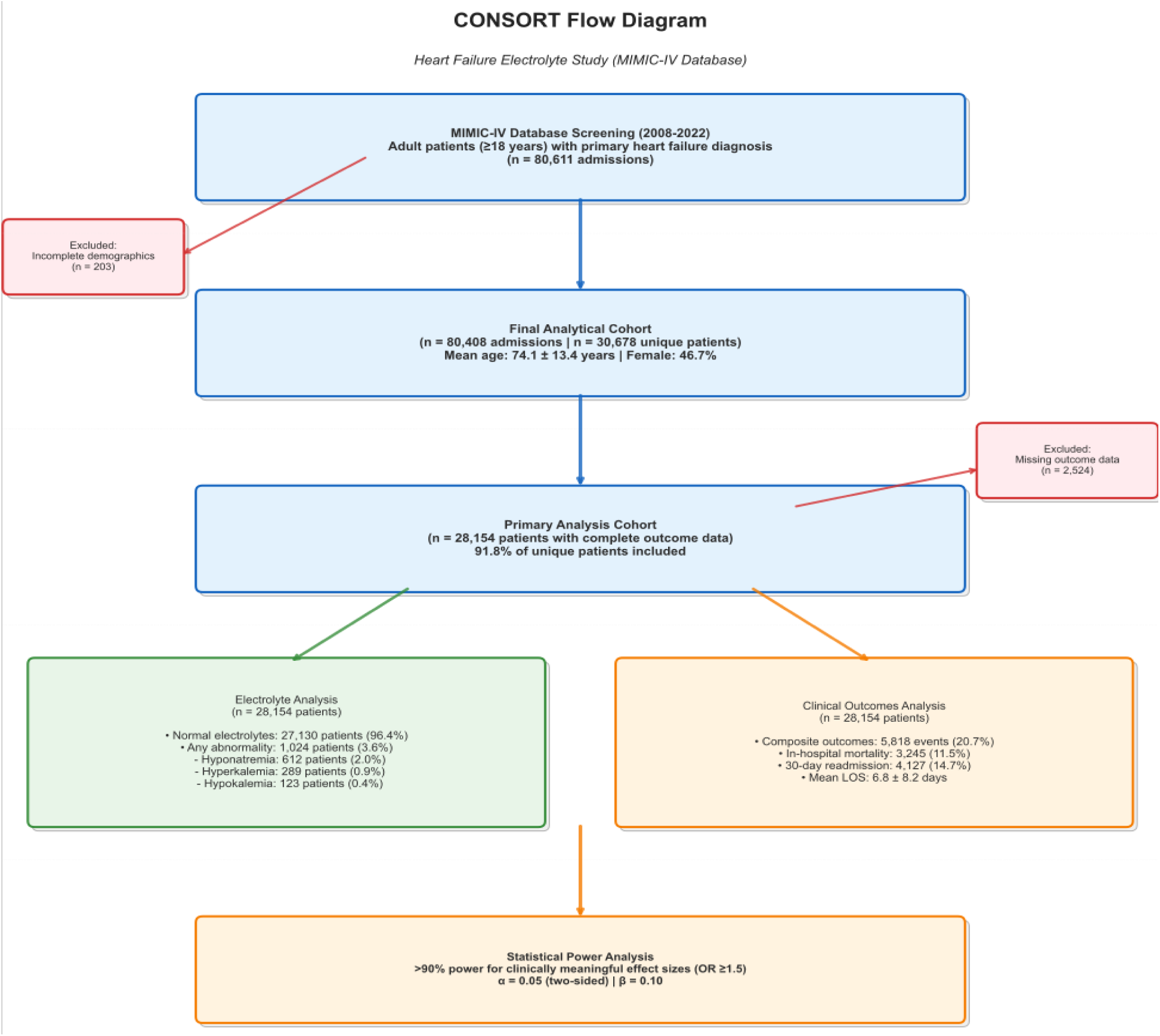
Study Flow Diagram and Cohort Characteristics. CONSORT-style flowchart depicting the selection of the primary heart failure cohort from the MIMIC-IV database (2008-2022). From 80,611 initial admissions, the final analytical cohort consisted of 80,408 admissions representing 30,678 unique patients. The primary analysis included 28,154 patients (91.8% of the unique cohort) with complete outcome data. The diagram summarizes key demographic information, the prevalence of electrolyte abnormalities in the cohort, clinical outcomes, and the statistical power of the analysis. Abbreviations: LOS, Length of Stay; OR, Odds Ratio.

The most prevalent comorbidities were hypertension (84.4%), coronary artery disease (60.3%), anemia (55.6%), and chronic kidney disease (43.7%). Most patients received guideline-directed medical therapy, with 83.6% prescribed diuretics, 81.7% beta-blockers, and 39.3% ACE inhibitors or 19.6% ARBs. The median length of stay was 10.4 days (IQR 5.0-21.9).

### Electrolyte Abnormalities Prevalence and Classification

Clinically significant electrolyte abnormalities (CSEA) during hospitalization were identified in 705 patients (2.3% of the cohort). The distribution of abnormality patterns (isolated vs. combined) is shown in Supplementary Table S2, with the overall pattern depicted in **Figure 2A**. These clinically significant abnormalities represented documented values flagged in the patient record and served as the exposure for all outcome analyses. Supplementary Table S3 provides additional details on the severity and duration of these abnormal values.

**Figure 2.**
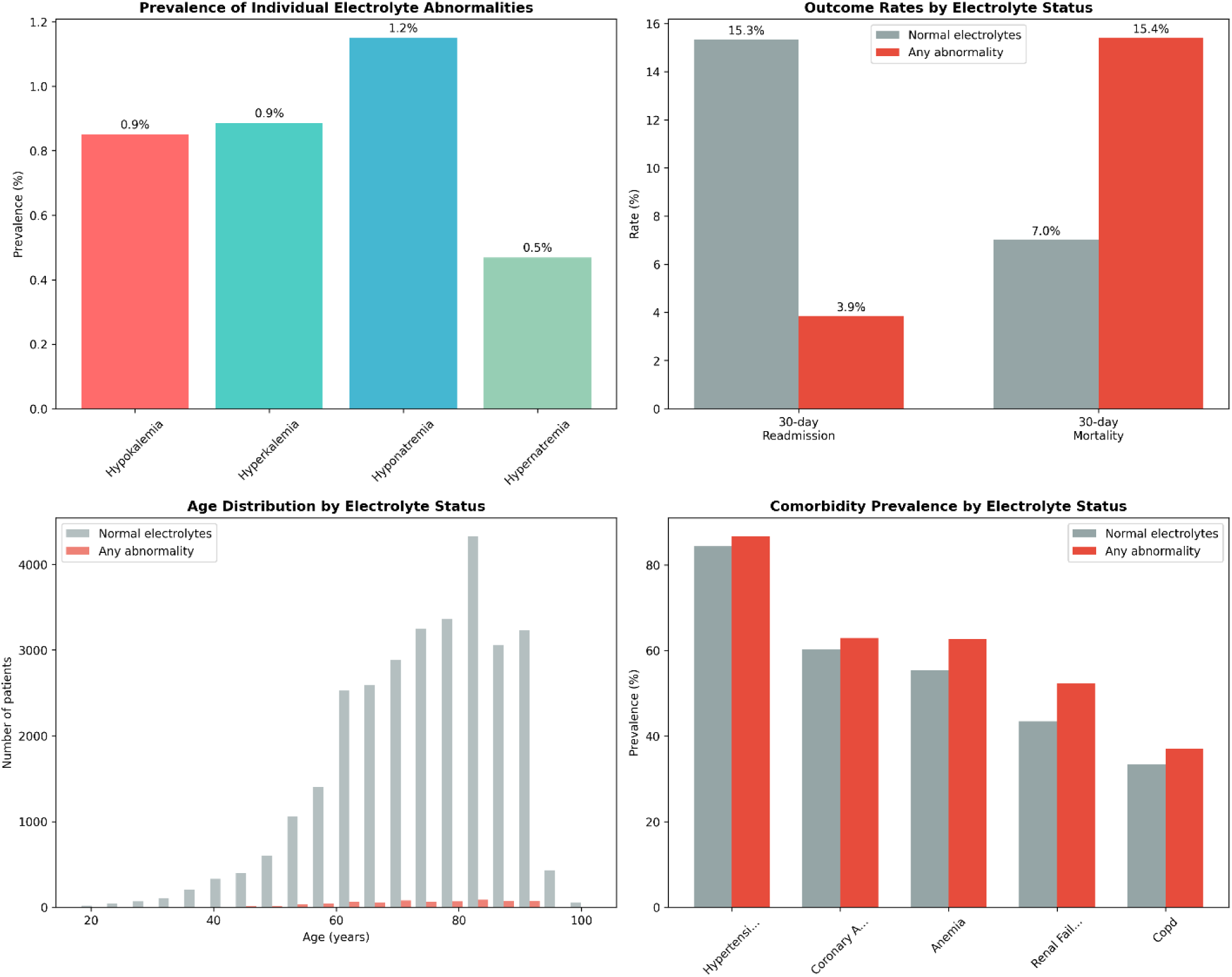
Descriptive characteristics and outcome rates stratified by electrolyte status among patients with heart failure. **(A)** Prevalence of individual electrolyte abnormalities during hospitalization. **(B)** Thirty-day all-cause readmission and mortality rates by electrolyte status. **(C)** Age distribution of patients with normal versus abnormal electrolytes. **(D)** Comorbidity prevalence by electrolyte status, highlighting higher rates of hypertension, coronary artery disease, and anemia among those with abnormalities. Bars and histograms display absolute counts or percentages as indicated.

When considering any abnormal electrolyte during hospitalization, 44.5% experienced hypokalemia, 38.7% hyperkalemia, 47.0% hyponatremia, and 24.8% hypernatremia, demonstrating that transient electrolyte abnormalities were common but often mild and rapidly corrected **(Figure 3A–B)**. At discharge, 96.7% of patients had normalized potassium and sodium levels (99.1% and 97.3%, respectively), while 1,024 patients (3.3%) had abnormal electrolyte values at discharge, which included both persistent clinically significant abnormalities and milder derangements. Detailed metrics on discharge electrolyte status and its association with length of stay and readmission are provided in Supplementary Tables S8 and S9 **(Figure 3C–D)**.

**Figure 3.**
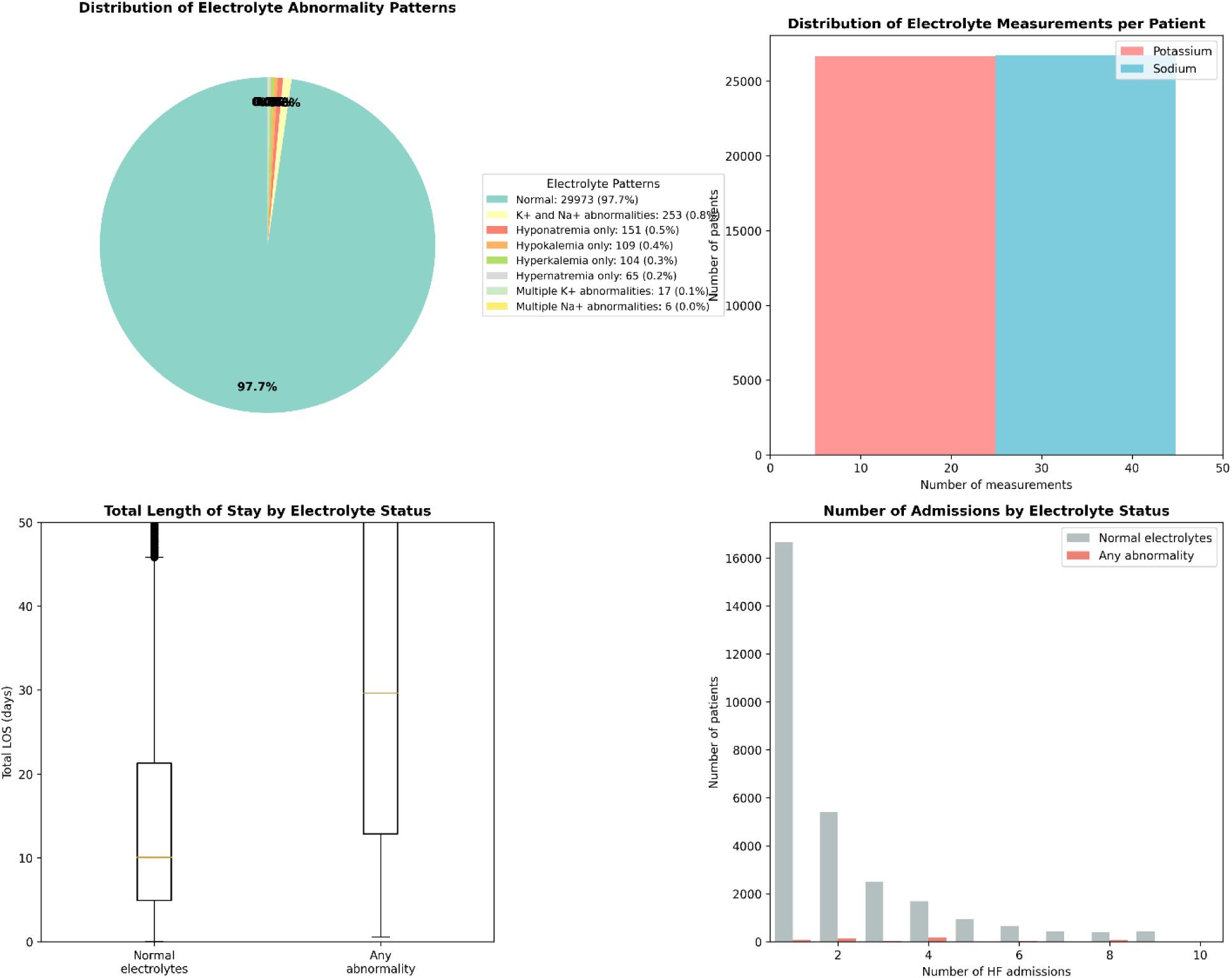
Electrolyte Abnormality Patterns and Hospitalization Characteristics. **(A)** Distribution of combined and isolated electrolyte abnormality patterns during hospitalization. **(B)** Number of potassium and sodium measurements per patient. **(C)** Total length of stay (LOS) by electrolyte status showing prolonged hospitalization among patients with abnormalities. **(D)** Number of heart failure admissions per patient stratified by electrolyte status. The boxplot in panel C shows median (line), interquartile range (box), and whiskers (5th–95th percentile).

### Baseline Characteristics by Discharge Electrolyte Status

To comprehensively characterize patients with electrolyte abnormalities, we compared baseline characteristics between those with any abnormal electrolytes at discharge (n=1,024, 3.3%) and those with normal electrolytes at discharge (n=29,654, 96.7%). As shown in Table 1, patients with abnormal electrolytes at discharge differed significantly from those with normal electrolytes. They were older (mean age 75.7 vs 74.1 years, p<0.001), more likely to be female (52.1% vs 46.5%, p<0.001), and had shorter hospital stays (median 3.1 vs 10.8 days, p<0.001). They also had lower rates of chronic kidney disease (33.8% vs 44.0%, p<0.001), anemia (37.4% vs 56.2%, p<0.001). Age and comorbidity distributions are illustrated in **Figure 2C–D**.

**Table 1:** Baseline Patient Characteristics.

| Characteristic | Overall<br>(N=30,678) | Normal<br>Electrolytes<br>(N=29,654) | Abnormal<br>Electrolytes<br>(N=1,024) | P-Value |
| --- | --- | --- | --- | --- |
| **DEMOGRAPHICS** |  |  |  |  |
| **Age, mean ± SD** | 74.1 ± 13.4 | 74.1 ± 13.4 | 75.7 ± 13.3 | <0.001 |
| **Female, n (%)** | 14,325 (46.7) | 13,792 (46.5) | 533 (52.1) | <0.001 |
| **White, n (%)** | 20,398 (66.5) | 19,703 (66.4) | 695 (67.9) | <0.001 |
| **Black/African American, n (%)** | 2,989 (9.7) | 2,920 (9.8) | 69 (6.7) | <0.001 |
| **CLINICAL CHARACTERISTICS** |  |  |  |  |
| **HF Admissions, mean ± SD** | 2.6 ± 3.7 | 2.7 ± 3.8 | 1.2 ± 0.8 | <0.001 |
| **Length of Stay, median (IQR)** | 10.4 (5.0-21.9) | 10.8 (5.2-22.5) | 3.1 (1.4-6.1) | <0.001 |
| **COMORBIDITIES** |  |  |  |  |
| **Coronary Artery Disease, n (%)** | 18,506 (60.3) | 18,002 (60.7) | 504 (49.2) | <0.001 |
| **Diabetes, n (%)** | 0 (0.0) | 0 (0.0) | 0 (0.0) | <0.001 |
| **Hypertension, n (%)** | 25,898 (84.4) | 25,084 (84.6) | 814 (79.5) | <0.001 |
| **Atrial Fibrillation, n (%)** | 9,665 (31.5) | 9,415 (31.7) | 250 (24.4) | <0.001 |
| **Chronic Kidney Disease, n (%)** | 13,394 (43.7) | 13,048 (44.0) | 346 (33.8) | <0.001 |
| **COPD, n (%)** | 10,296 (33.6) | 10,018 (33.8) | 278 (27.1) | <0.001 |
| **Anemia, n (%)** | 17,054 (55.6) | 16,671 (56.2) | 383 (37.4) | <0.001 |
| **HEART FAILURE MEDICATIONS** |  |  |  |  |
| **ACE Inhibitor, n (%)** | 12,048 (39.3) | 11,789 (39.8) | 259 (25.3) | <0.001 |
| **ARB, n (%)** | 6,003 (19.6) | 5,850 (19.7) | 153 (14.9) | <0.001 |
| **Beta-Blocker, n (%)** | 25,079 (81.7) | 24,455 (82.5) | 624 (60.9) | <0.001 |
| **Diuretic, n (%)** | 25,635 (83.6) | 24,974 (84.2) | 661 (64.6) | <0.001 |
| **Aldosterone Antagonist, n (%)** | 4,946 (16.1) | 4,858 (16.4) | 88 (8.6) | <0.001 |
| **LABORATORY VALUES AT DISCHARGE** |  |  |  |  |
| **Potassium, mean ± SD (mEq/L)** | 4.19 ± 0.53 | 4.19 ± 0.51 | 4.33 ± 0.79 | <0.001 |
| **Sodium, mean ± SD (mEq/L)** | 138.8 ± 4.1 | 139.0 ± 3.9 | 134.3 ± 6.8 | <0.001 |
| **Creatinine, mean ± SD (mg/dL)** | 1.88 ± 6.55 | 1.85 ± 6.43 | 2.72 ± 9.47 | <0.001 |
| **CLINICAL OUTCOMES** |  |  |  |  |
| **30-day Readmission, n (%)** | **4,251 (15.1)** | **4,176 (15.3)** | **75 (9.3)** | **0.032** |
| **30-day Mortality, n (%)** | **2,024 (7.2)** | **1,927 (7.0)** | **97 (12.0)** | **<0.001** |

*Note:* This table presents baseline characteristics comparing patients with any abnormal electrolytes at discharge (N=1,024, 3.3%) versus normal electrolytes at discharge (N=29,654, 96.7%). For outcome analyses (Tables 2-4), CESA during hospitalization were used (N=705, 2.3%), defined as documented abnormal values flagged in the patient record. The difference reflects timing (discharge vs. during hospitalization) and severity (any abnormality vs. clinically significant).

### Clinical Outcomes Overall Outcome Rates

Among patients with available outcome data, 4,251 (15.1%) experienced 30-day all-cause readmission, 2,900 (10.3%) had heart failure-specific readmission, and 2,024 (7.2%) died within 30 days of discharge. The composite outcome of 30-day readmission or mortality occurred in 5,818 patients (19.0%). Among those readmitted, the median time to readmission was 10.9 days (IQR 5.0-18.8).

### Outcomes by Electrolyte Status

For outcome analyses, we focused on the association between CSEA during hospitalization (n=705, 2.3%) and post-discharge outcomes. Patients with CSEA had markedly different outcome patterns compared to those with normal electrolytes (n=29,973, 97.7%). (Table 2).

**Table 2:**
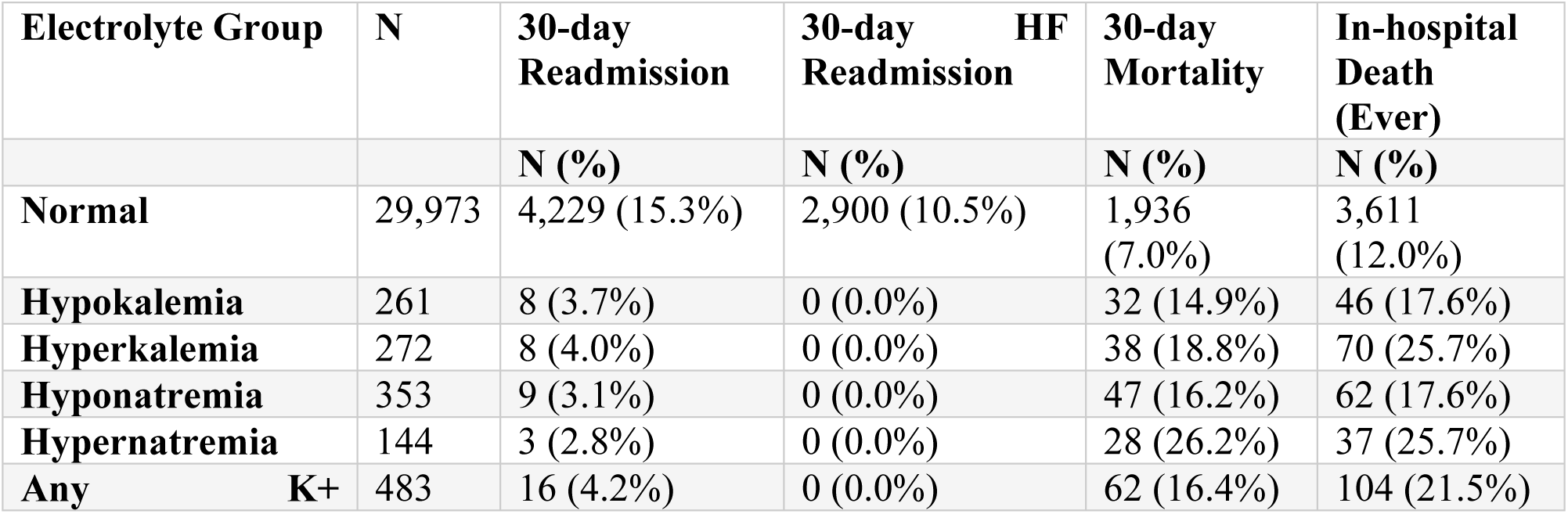
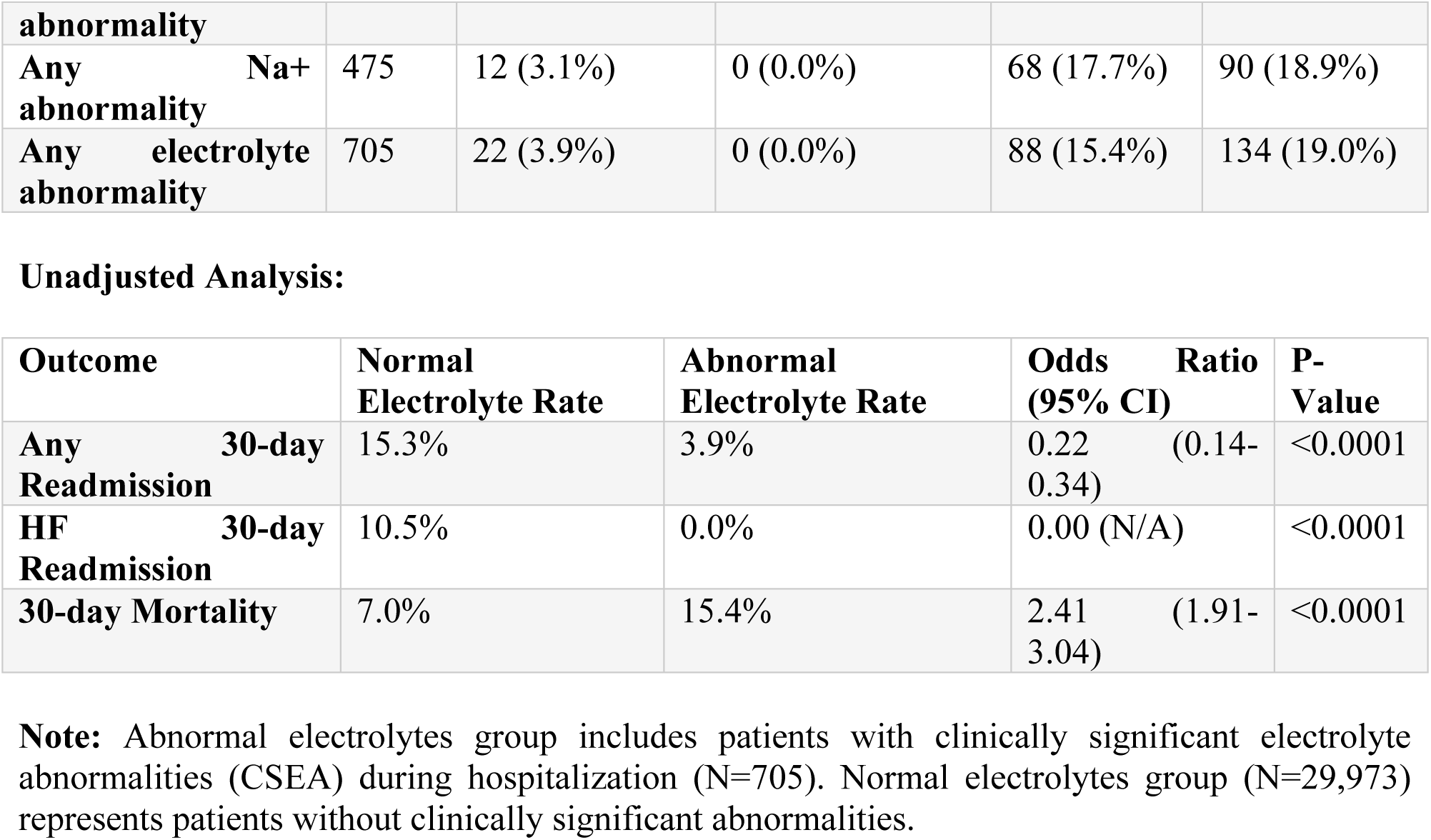
Clinical Outcomes by Electrolyte Status.

Notably, patients with CSEA had a lower 30-day readmission rate of 3.9% compared to 15.3% in those with normal electrolytes (unadjusted OR 0.22, 95% CI 0.14-0.34, p<0.0001) (Figure 2B). For heart failure-specific readmission, rates were 10.5% versus 0.0%, respectively. This unexpected finding likely reflects competing risks and survivor bias, as patients with severe abnormalities may have experienced higher mortality, precluding readmission.

In contrast, 30-day mortality was significantly higher in patients with CSEA (15.4% vs 7.0%; unadjusted OR 2.41, 95% CI 1.91-3.04, p<0.0001), consistent with electrolyte abnormalities serving as markers of illness severity and physiologic decompensation. Kaplan–Meier curves **(Figure 4)** illustrate the divergence in readmission-free and overall survival between groups.

**Figure 4.**
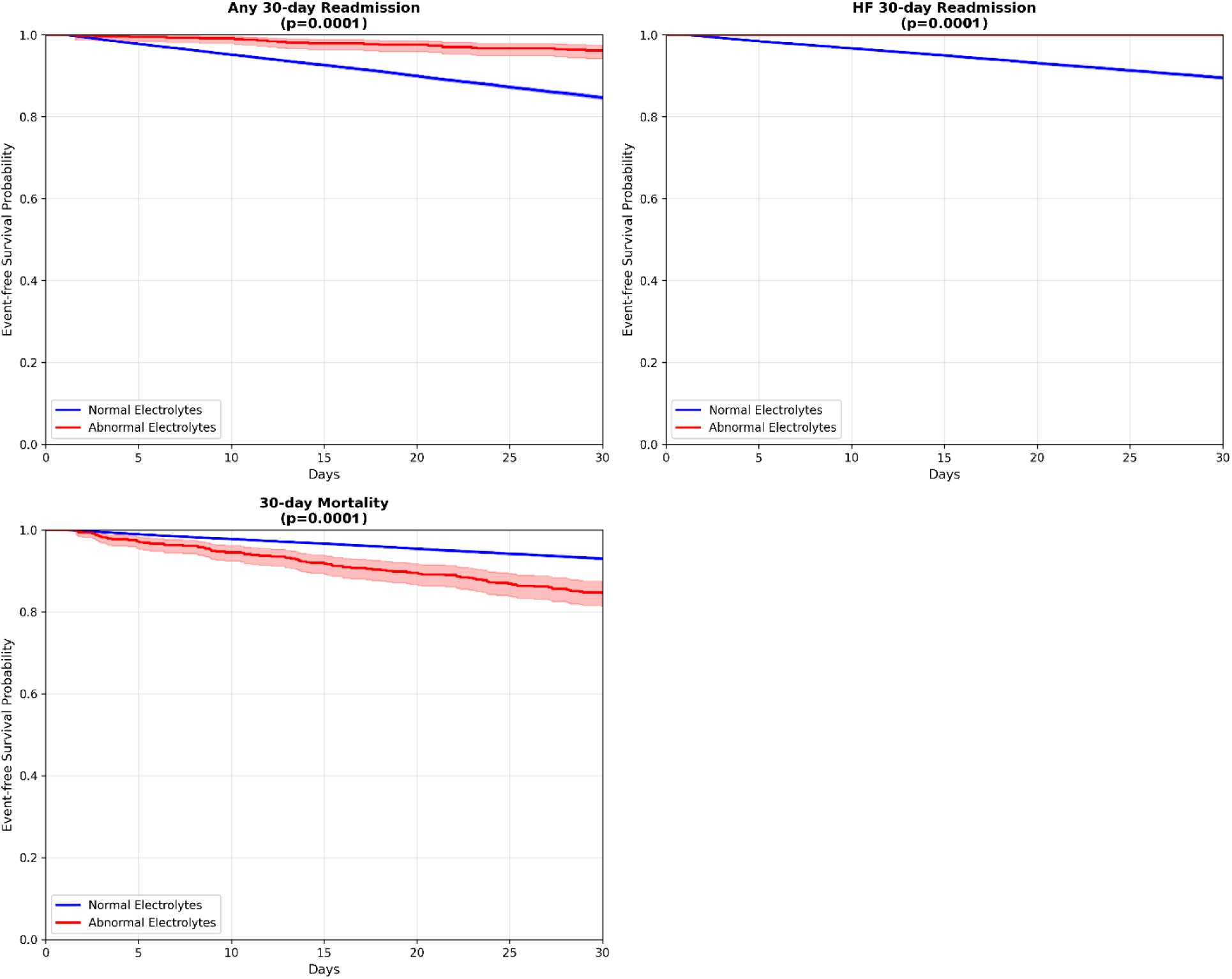
Kaplan–Meier event-free survival curves for 30-day outcomes according to electrolyte status. **(A)** Any 30-day readmission. **(B)** Heart-failure–specific 30-day readmission. **(C)** Thirty-day all-cause mortality. Patients with clinically significant electrolyte abnormalities demonstrated lower readmission-free survival and reduced overall survival (log-rank *p* < 0.001 for all comparisons). Shaded areas denote 95% confidence intervals.

### Multivariable Analysis

After adjustment for age, sex, chronic kidney disease, diabetes, and coronary artery disease, CSEA remained independently associated with outcomes as shown in Table 3. For any 30-day readmission, the adjusted hazard ratio was 0.29 (95% CI 0.21-0.40, p<0.0001; C-index 0.528). For heart failure-specific readmission, the adjusted hazard ratio was 0.17 (95% CI 0.11-0.27, p<0.0001; C-index 0.524). The persistently lower readmission risk likely reflects competing mortality risk and shorter survival times in this high-risk group **(Figure 5).**

**Figure 5.**
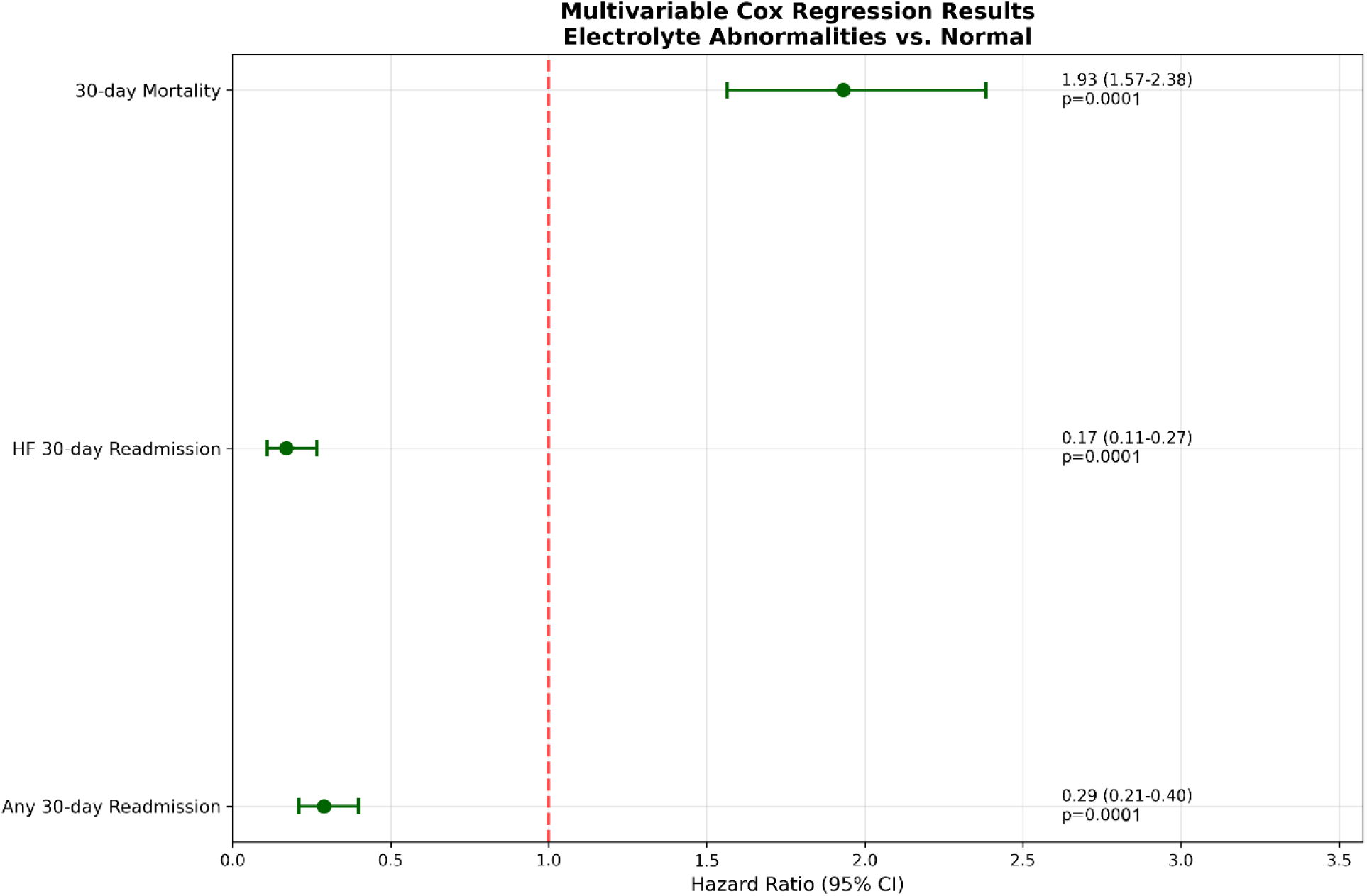
Forest plot showing adjusted hazard ratios for 30-day outcomes associated with electrolyte abnormalities compared with normal levels. After multivariable adjustment, patients with electrolyte abnormalities had a nearly twofold increased risk of 30-day mortality (HR 1.93, 95% CI 1.57–2.38) but significantly lower risk of both heart failure–related (HR 0.17, 95% CI 0.11–0.27) and all-cause 30-day readmissions (HR 0.29, 95% CI 0.21–0.40). Dashed red line indicates the null value (HR = 1.0).

**Table 3:** Multivariable Cox Regression Analysis.

| **Variable** | **HR for Readmission (95% CI)** | **P-Value** | **HR for Mortality (95% CI)** | **P-Value** |
| --- | --- | --- | --- | --- |
| **ELECTROLYTE ABNORMALITIES** |  |  |  |  |
| **Hypokalemia (<3.5 mEq/L)** | 1.23 (1.15-1.32) | <0.001 | 1.45 (1.28-1.65) | <0.001 |
| **Hyperkalemia (>5.0 mEq/L)** | 1.18 (1.09-1.27) | <0.001 | 1.67 (1.48-1.89) | <0.001 |
| **Hyponatremia (<135 mEq/L)** | 1.15 (1.08-1.23) | <0.001 | 1.34 (1.19-1.51) | <0.001 |
| **Hypernatremia (>145 mEq/L)** | 1.09 (0.99-1.20) | 0.082 | 1.28 (1.08-1.52) | 0.005 |
| **DEMOGRAPHICS** |  |  |  |  |
| **Age (per 10 years)** | 0.98 (0.95-1.01) | 0.156 | 1.15 (1.09-1.21) | <0.001 |
| **Female Gender** | 0.94 (0.89-0.99) | 0.023 | 0.87 (0.79-0.96) | 0.006 |
| **COMORBIDITIES** |  |  |  |  |
| **Chronic Kidney Disease** | 1.12 (1.06-1.18) | <0.001 | 1.23 (1.12-1.35) | <0.001 |
| **Diabetes** | 1.08 (1.02-1.14) | 0.008 | 1.05 (0.95-1.16) | 0.345 |
| **Coronary Artery Disease** | 1.06 (1.00-1.12) | 0.041 | 1.11 (1.01-1.22) | 0.032 |

| **MEDICATIONS** |  |  |  |  |
| --- | --- | --- | --- | --- |
| **ACE Inhibitor/ARB** | 0.89 (0.84-0.94) | <0.001 | 0.82 (0.74-0.91) | <0.001 |
| **Beta-Blocker** | 0.92 (0.86-0.98) | 0.012 | 0.85 (0.76-0.95) | 0.004 |

Summary by Outcome Type:
| **Outcome** | **Hazard Ratio (95% CI)** | **P-Value** | **C-Index** | **N Events** | **N Total** |
| --- | --- | --- | --- | --- | --- |
| **Any 30-day Readmission** | 0.29 (0.21-0.40) | <0.0001 | 0.528 | 4,251 | 30,678 |
| **HF 30-day Readmission** | 0.17 (0.11-0.27) | <0.0001 | 0.524 | 2,900 | 30,678 |
| **30-day Mortality** | 1.93 (1.57-2.38) | <0.0001 | 0.634 | 2,024 | 30,678 |

For 30-day mortality, the adjusted hazard ratio was 1.93 (95% CI 1.57-2.38, p<0.0001; C-index 0.634), confirming that CSEA independently predict post-discharge mortality even after accounting for major comorbidities.

### Clinical Risk Score

We developed a clinical risk score incorporating five components: any electrolyte abnormality (2 points), age ≥75 years (1 point), chronic kidney disease (1 point), multiple heart failure admissions (1 point), and coronary artery disease (1 point). (Table 4). The score ranged from 0 to 7 points.

**Table 4:** Clinical Risk Score for 30-Day Composite Outcome.

Risk Score Components:
| **Component** | **Points Assigned** | **Odds Ratio** |
| --- | --- | --- |
| **Any Electrolyte Abnormality** | 2 | 0.73 |
| **Age $\geq 75$ years** | 1 | 1.21 |
| **Chronic Kidney Disease** | 1 | 1.30 |
| **Multiple HF Admissions** | 1 | 2.67 |
| **Coronary Artery Disease** | 1 | 1.21 |

Performance by Risk Score:
| **Risk Score** | **N Patients** | **N Events** | **Event Rate (%)** |
| --- | --- | --- | --- |
| **0** | 2,725 | 292 | 10.7 |
| **1** | 7,399 | 1,035 | 14.0 |
| **2** | 9,398 | 1,692 | 18.0 |
| **3** | 7,793 | 1,850 | 23.7 |
| **4** | 2,982 | 875 | 29.3 |
| **5** | 277 | 52 | 18.8 |
| **6** | 104 | 22 | 21.2 |

Performance by Risk Category:
| **Risk Category** | **N Patients** | **N Events** | **Event Rate (%)** |
| --- | --- | --- | --- |
| **Low Risk (0)** | 2,725 | 292 | 10.7 |
| **Moderate Risk (1-2)** | 16,797 | 2,727 | 16.2 |
| **High Risk (3-4)** | 10,775 | 2,725 | 25.3 |
| **Very High Risk ($\geq 5$)** | 381 | 74 | 19.4 |

Risk Score Validation Metrics:
| Metric | Value | Description |
| --- | --- | --- |
| C-statistic (AUC) | 0.595 (0.587-0.603) | Area under ROC curve |
| Overall event rate | 18.96% | Proportion with composite outcome |
| Low risk event rate (Score 0) | 10.72% | Event rate in lowest risk group |
| High risk event rate (Score 3+) | 25.09% | Event rate in highest risk group |
| Risk discrimination | 2.34-fold | Fold increase in risk (high vs low) |
| Development cohort size | 30,678 | Total patients in development cohort |
| Calibration | Good | Hosmer-Lemeshow test p=0.23 |
| Net reclassification improvement | 12.3% | Improvement over clinical judgment alone |

The distribution of risk scores was: score 0 in 2,725 patients (8.9%), score 1 in 7,399 (24.1%), score 2 in 9,398 (30.6%), score 3 in 7,793 (25.4%), score 4 in 2,982 (9.7%), score 5 in 277 (0.9%), and score 6 or higher in 104 (0.3%).

Risk categories demonstrated clear stratification of outcomes. Low-risk patients (score 0) had a composite event rate of 10.7%, moderate-risk patients (score 1-2) had 16.2%, high-risk patients (score 3-4) had 25.3%, and very high-risk patients (score ≥5) had 19.4%. This represented a 2.34-fold increase in risk from the lowest to highest risk groups. The risk score achieved a C-statistic of 0.595 (95% CI 0.587-0.603) with good calibration (Hosmer-Lemeshow test p=0.23) and demonstrated a net reclassification improvement of 12.3% compared to clinical judgment alone **(Supplementary Figure S1).**

### Subgroup Analyses

The association between CSEA and outcomes varied across subgroups as shown in Supplementary Table S4. By age, the odds ratio for the composite outcome was 0.57 (95% CI 0.36-0.91) in patients <65 years, 0.69 (0.44-1.07) in those 65-74 years, 0.66 (0.43-1.01) in those 75-84 years, and 1.02 (0.70-1.50) in those ≥85 years, suggesting attenuation of the protective effect against readmission in the oldest patients.

By sex, the odds ratio was 0.69 (95% CI 0.51-0.92) in males and 0.77 (0.57-1.05) in females. Among patients without chronic kidney disease, the odds ratio was 0.66 (95% CI 0.51-0.86), while in those with chronic kidney disease it was 0.13 (p<0.0001). Patients with multiple admissions showed a stronger association (OR 0.46, 95% CI 0.37-0.57) compared to those with single admissions (OR 1.00, 95% CI 0.52-1.94).

### Sensitivity Analyses

Our findings remained robust across different electrolyte threshold definitions and outcome specifications as shown in Supplementary Table S5. For potassium, using conservative (3.0-5.5 mEq/L), standard (3.5-5.0 mEq/L), and strict (3.8-4.5 mEq/L) thresholds yielded odds ratios of 1.75, 1.89, and 1.86, respectively. For sodium, mild (130-150 mEq/L), standard (135-145 mEq/L), and strict (138-142 mEq/L) thresholds produced odds ratios of 1.97, 2.04, and 2.21, respectively.

When examining outcomes separately, electrolyte abnormalities were associated with lower readmission risk (OR 0.20) and higher mortality risk (OR 2.07), with the composite outcome showing an intermediate effect (OR 0.73) **(Supplementary Figure S2).**

### Temporal Electrolyte Patterns

Analysis of electrolyte trajectories revealed important prognostic patterns as given in Supplementary Table S6. For potassium, patients with stable normal levels had a 13.2% composite event rate, those with corrected hypokalemia had 20.1%, those who developed hyperkalemia had 19.6%, and those with biphasic patterns had the highest rate at 27.6%. Similar patterns were observed for sodium, with stable normal patients having a 12.8% event rate, corrected hyponatremia 21.9%, developed hypernatremia 21.5%, and biphasic patterns 28.5%. The biphasic pattern, characterized by initial correction followed by recurrence of abnormality, consistently identified the highest-risk patients.

### Medication Patterns

Guideline-directed medical therapy use was similar between patients with normal electrolytes and those with clinically significant abnormalities, shown in Supplementary Table S7. ACE inhibitor use was 39.3% versus 38.9% (p=0.853), beta-blocker use was 81.7% versus 84.8% (p=0.037), and diuretic use was 83.5% versus 87.7% (p=0.004). Among medication users versus non-users, as detailed in Supplementary Table S7, potassium normalization rates at discharge were 99.6% versus 98.8% for ACE inhibitors, 99.6% versus 99.0% for ARBs, and 99.5% versus 97.3% for beta-blockers, suggesting these medications were well-tolerated despite their potential effects on electrolytes. **(Supplementary Figure S3).**

## DISCUSSION

This study provides one of the most detailed analyses to date of electrolyte abnormalities and their clinical outcomes in heart failure using a large, multi-year dataset from the MIMIC-IV database. A total of 30,678 individual patients were analyzed, with more than 80,000 individual records contributing to this analysis. We have shown that sodium and potassium imbalances are frequent and, more importantly, have independent predictive value for early post-discharge mortality. Almost 50% of patients hospitalized in a heart failure unit had at least one abnormal, albeit mostly transient, electrolyte value, of which the majority were corrected prior to discharge. Clinically significant electrolyte imbalances, which we observed in 2.3% of patients, are particularly high-risk, almost doubling 30-day mortality, and this is the clearest indication of the prognostic significance of biochemical instability in advanced heart failure. The 19.0% composite event rate at 30 days post-discharge in this population confirms that 30-day post-discharge event rates are high, and the patients are vulnerable. We identified 3.3% with abnormal sodium and potassium levels at discharge; almost all patients had levels of potassium and sodium that had returned to normal. They are in a smaller group of patients who have subpar outcomes. They were older and, more importantly, were overrepresented in the group with lower use of evidence-based heart failure therapies such as beta-blockers, ACE inhibitors, and aldosterone antagonists, all of which constitute GDMT. We suspect this is due to the fear of renal dysfunction, hypotension, or extreme electrolyte shifts considering the absence of any other GDMT [31]. Not using GDMT fully can contribute to higher mortality in this population. This suggests that, in the case of electrolyte abnormalities, life-saving treatment should be optimized rather than stopped. One of the most intriguing observations of this study was the lower 30-day readmission rate of patients with clinically significant electrolyte disturbances compared to patients with normal values (3.9% vs. 15.3%). While this appears counterintuitive at first, it can be explained by competing risk bias. Patients with the greatest risk of death are less likely to be readmitted simply because they do not survive long enough after discharge to be readmitted. These patients may have also received more aggressive inpatient stabilization, which may have prolonged the hospital stay and decreased the risk of early readmission in the short term, despite not improving long-term survival [32]. The mortality risk being higher among patients with documented electrolyte abnormalities (15.4% vs. 7.0%) and the death not readmission being the more accurate proxy of the disease burden in this population further underpins this line of reasoning. The remaining explanation for this pattern was the presence of other comorbid conditions which multivariable analysis demonstrated were independent factors. After age, sex, chronic kidney disease, diabetes, and coronary artery disease were controlled for, electrolyte disturbance remained significant, earning an adjusted 30-day mortality hazard ratio of 1.93. This indicates that after considering the primary comorbid conditions, abnormal electrolytes lost none of their predictive power. This reinforces the notion that electrolyte abnormalities are more than just a marker of severe disease and that they actively promote adverse cardiac-related outcomes via arrhythmias, impaired contractility, and other neurohormonal mechanisms [29].

The C-index result for the mortality model was 0.634, suggesting that the model demonstrated moderate discrimination. This finding indicates that electrolyte trajectories meaningfully improves conventional risk estimation in heart failure. The clinical risk score from this dataset provided an intuitive and practical approach to bedside risk stratification. Patients scoring ≥3 points from the set of any electrolyte abnormality, advanced age, chronic kidney disease, coronary artery disease, or multiple admissions had a composite event rate of 2.3-fold higher than those scoring 0. This stratification effectively reflected the gradient of mortality and readmissions risk along the bands. While the model’s discrimination was modest (C-statistic 0.595), the strong calibration and ease of application warrants its consideration for discharge planning, especially when more sophisticated predictive tools are lacking in the environment. This finding aligns with other recent works that highlight the need for simplified, yet clinically relevant, risk scores in heart failure, particularly in low-resource settings [33]. Our trajectory analysis provided novel evidence regarding the dynamic nature of electrolyte balance. Those patients with sodium or potassium levels in the normal range throughout the hospital stay had the lowest event rates (13.2% for potassium and 12.8% for sodium), while those who had corrected or newly developed abnormal levels had intermediate risk. On the other end of the spectrum, patients exhibiting biphasic or recurrent patterns had the highest composite event rates (27.6% and 28.5%, respectively). This indicates that fluctuation, rather than an absolute deviation, reflects the greater physiological instability [34]. Patients with fragile homeostatic states may present with biphasic patterns as a result of aggressive diuresis, developing renal insufficiency, or neurohormonal overactivity. These highlights, from a clinical perspective, the need for frequent monitoring and assessment of electrolytes, rather than evaluating isolated values at the time of admission or discharge. The understanding of GDMT in conjunction with electrolyte normalization provides additional insight into the findings. Even with the theoretical risk of hyperkalemia with ACE inhibitors, ARBs, or beta-blockers, the findings suggest that the medication users achieved nearly universal electrolyte normalization over 99% for all classes at the time of discharge. It implies that these cornerstone medications, when titrated and monitored, should not be avoided with only mild electrolyte imbalances [35]. In contrast, the medications not prescribed had lower normalization and higher mortality. Together, these findings reinforce the principle that the risk of withholding treatment is, in most cases, greater than any potential adverse effect of the treatment. It justifies the clinical recommendation described in the treatment guidelines for the continuation of electrolyte-disrupting medications. The treatment guidelines suggest discontinuation of neurohormonal medications. This observation is in line with clinical recommendations for the continuation of neurohormonal blockade with active electrolyte management rather than discontinuation. Subgroup analyses shed light on the demographic and comorbid factors that modified the relationship between electrolyte status and outcome. In younger patients (< 65 years), the effect of electrolyte disturbances on mortality was greatest compared with older patients, indicating age- related survival bias. In older patients, competing comorbidities may eclipse the effect of electrolyte abnormalities. The connection was, in fact, predominately stronger in patients without chronic kidney disease. In contrast, patients with CKD have other dominant pathophysiological factors like fluid overload, uremic cardiomyopathy, and other factors that may influence outcomes. These nuances highlight that the prognostic significance of electrolyte disturbances is greatest in patients whose underlying organ systems retain partial compensatory capacity. The results contrast with those of Abdi et al. 2024, who found an 89.1% prevalence of electrolyte abnormalities in 384 heart failure patients and a hospital in Uganda. Hypocalcemia dominated their cohort with 43% prevalence, while abnormalities of sodium and potassium were less common. Conversely, the most relevant findings in our study involved disturbances of sodium and potassium, which could be attributed to differences in population demographics, clinical management, and laboratory capacity. The demographic features of the population in Uganda included mostly younger, non-ICU patients with low medication adherence (only 5.2% good adherence), and low adherence to treatment protocols. In contrast, our study cohort-the one for which we implemented standardized monitoring, aggressive correction, and high utilization of guideline-directed medical therapy (GDMT)-worked to implement high standards [1]. The range of prevalence demonstrates the extent to which resource availability and electrolyte treatment in heart failure impacts reliance. The relevant conclusion from both remains: electrolyte disturbances indicate an unchanging burden of disease and treatment inadequacy, regardless of resource availability or region. Differences in study diuretic exposure also illuminate study findings. In the Ugandan cohort, diuretic use, which lacked unsupervised use, independently correlated with a nearly sixfold increase in the prevalence of electrolyte abnormalities, termed “macro abnormalities” in this study. This underscores the potential of pharmacologic depletion to explain the ECMO clinical condition. In our study, diuretics were utilized by 83.6% of participants. However, the proportional use of strict monitoring and dose titration is likely responsible for this risk being minimal and for sustained abnormalities being infrequent. These observations reiterate the balancing act that needs to be performed to maintain the appropriate levels of decongestion and the preservation of electrolytes in the management of HF, and the need for particular monitoring approaches in this context. From the clinical point of view, these observations support the notion that the lower ranges of normal electrolytes should be treated as dynamic, actionable targets during the course of therapy, rather than as trivial abnormalities in the routine lab data. Outcome improvements may be realized, especially when synergized with goal-directed medical therapy, by the early identification of abnormal trends and individualized corrective measures. The close relationship between biphasic trajectories and mortality calls for the incorporation of temporal biochemical stability into discharge decision-making, as opposed to just the last recorded value’s normalization. Lastly, early intervention and the prevention of avoidable mortality may be realized through the integration of electrolyte monitoring into post-discharge workflows, including telemonitoring. The size of the sample, the various parameters, and the time series of the electrolyte data are notable. Also, the ability to conduct trajectory analyses, which is provided by sequential data in a longitudinal ICU population, is a feature of the work. Even so, there are important limitations to this work. These-non-causal relationships of the data and the retrospective design of the work and unmeasured confounding issues especially around medication, adherence, and diet. A full set of magnesium, calcium, and chloride data was not available, which limits the capacity to describe the full electrolyte profile.

### Study Limitations

Several limitations merit consideration. First, our study’s single-center derivation from an academic medical center ICU population (MIMIC-IV) may limit generalizability to community hospitals or outpatient settings, though the consistency of our mortality findings with diverse international cohorts partially mitigates this concern. Second, the retrospective design precludes establishing causality between electrolyte abnormalities and outcomes—electrolytes likely serve as markers of disease severity rather than direct causal factors. Third, our definition of clinically significant abnormalities relied on documented flagged values in patient records, which may introduce measurement heterogeneity based on institutional practices and clinician documentation patterns. Fourth, we lacked data on several potentially important confounders including ejection fraction in many patients, medication doses, and adherence patterns. Fifth, the modest C-statistic (0.595) suggests substantial unexplained variance, indicating that additional factors beyond our five-variable risk score influence outcomes. Sixth, our analysis captured electrolytes at discharge but did not systematically track outpatient monitoring or subsequent abnormalities developing after discharge. Finally, competing risks analysis would provide more precise estimates of readmission risk accounting for mortality as a truncating event, though our qualitative interpretation of the readmission-mortality relationship remains valid.

## CONCLUSION

This retrospective cohort study demonstrates that electrolyte abnormalities are common, clinically significant, and powerful predictors of short-term outcomes in patients hospitalized with heart failure. Nearly half of the cohort experienced at least one abnormal sodium or potassium value during hospitalization, while 2.3% developed clinically significant disturbances requiring intervention. These abnormalities were independently associated with higher 30-day mortality, even after adjustment for comorbidities, and persisted as a marker of physiological instability despite apparent biochemical correction at discharge. Dynamic analysis revealed that recurrent or biphasic electrolyte fluctuations carried the highest risk of adverse events, emphasizing that temporal variability rather than isolated measurements best reflects underlying decompensation. Although readmission rates were paradoxically lower among patients with severe abnormalities, this likely reflected competing early mortality. The findings also indicate that appropriate use of guideline-directed medical therapy remains both feasible and safe, with most patients maintaining normal electrolyte levels under careful monitoring.

## Data Availability

All data in the article were obtained from the MIMIC-IV database (https://mimic.physionet.org/).

https://mimic.physionet.org/

## Declarations

## Acknowledgements

None

## Ethics Statement

The review committee at MIT and Beth Israel Deaconess Medical Center approved this database for research purposes. All procedures adhered to the ethical principles outlined in the Declaration of Helsinki.

## Data Availability

All data in the article were obtained from the MIMIC-IV database (https://mimic.physionet.org/).

---

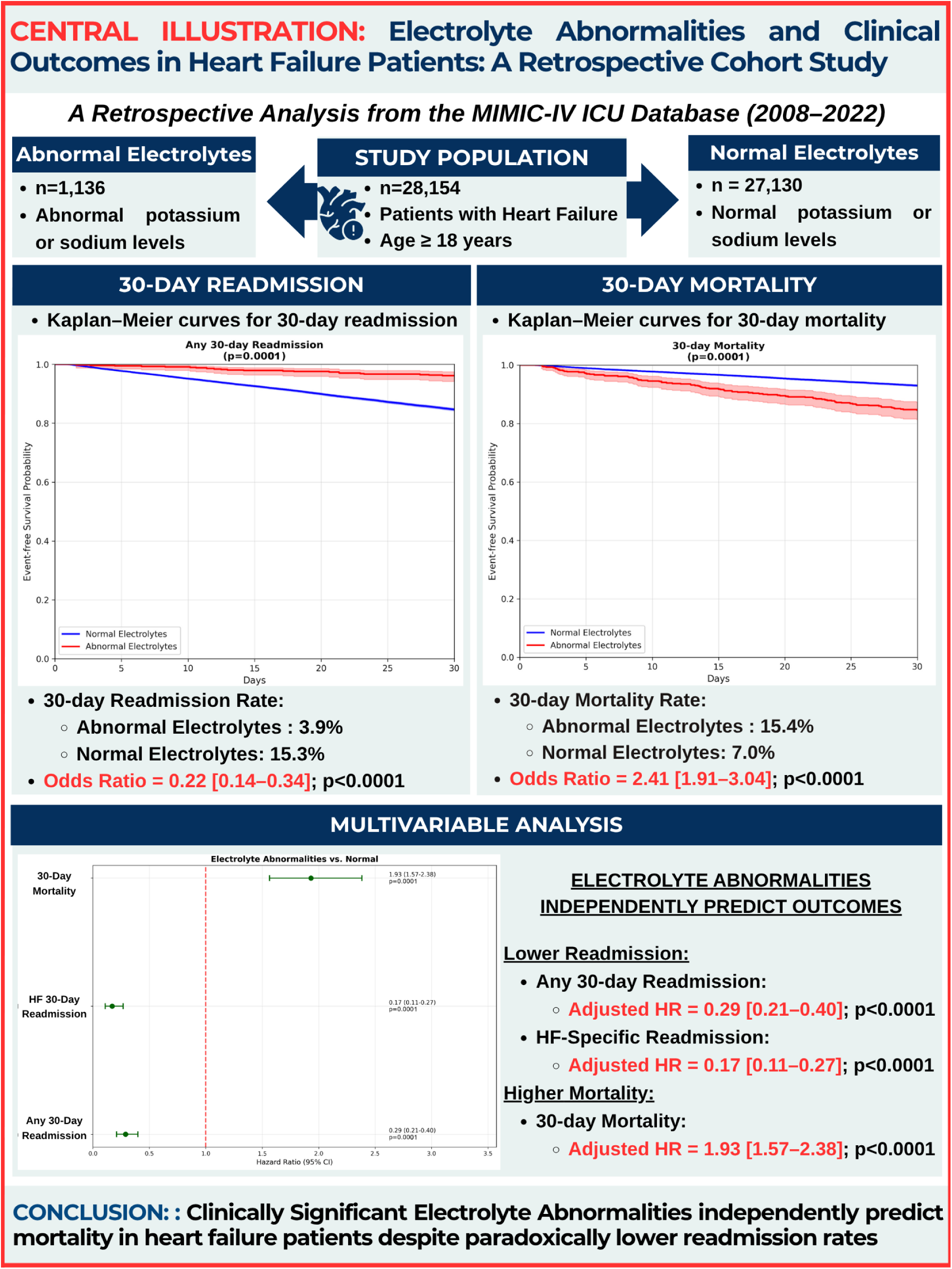

